# Role of exercise prehabilitation on surgical prognosis in people with breast cancer: A Systematic review and call for action

**DOI:** 10.1101/2025.10.29.25338895

**Authors:** Alba Esteban-Simón, Pablo Corres, Alberto Soriano-Maldonado, Almudena Carneiro-Barrera, Francisco J. Amaro-Gahete

**Affiliations:** Department of Education, Faculty of Education Sciences, University of Almería, Almería, Spain. CIBIS Research Centre, SPORT Research Group (CTS-1024), University of Almería, 04120 Almería, Spain; GIzartea, Kirola eta Ariketa Fisikoa Ikerkuntza Taldea (GIKAFIT). Society, Sports, and Exercise Research Group. Department of Physical Education and Sport, Faculty of Education and Sport, Physical Activity and Sport Sciences Section, University of the Basque Country (UPV/EHU), Vitoria-Gasteiz, Basque Country, Spain. Physical Activity, Exercise and Health Group, Bioaraba Health Research Institute, 01009 Vitoria-Gasteiz, Spain; Department of Psychology, Universidad Loyola Andalucía, 41704 Seville, Spain; Department of Physiology, Faculty of Medicine, University of Granada, Granada, Spain. CIBER de Fisiopatología de la Obesidad y Nutrición (CIBEROBN), Instituto de Salud Carlos III, Granada, Spain. Instituto de Investigación Biosanitaria, Ibs. Granada, Granada, Spain

**Keywords:** breast surgery, exercise, nutrition, psychological support

## Abstract

**Background:** Breast cancer (BC) is the most common cancer worldwide, with surgery being the main curative approach. However, postoperative complications (PC) are frequent and may compromise recovery. Identifying strategies to optimize surgical prognosis is clinically relevant.

**Objective:** To synthesize the effects of multimodal, exercise-based prehabilitation on PC, hospital length of stay (HLS), reoperations, and readmissions in patients with BC undergoing surgery.

**Methods:** A systematic search was conducted in PubMed and Web of Science. Eligible studies were randomized or non-randomized controlled trials including patients with BC, implementing prehabilitation with physical activity or exercise, and assessing PC, HLS, reoperations, and/or readmissions.

**Results:** Two studies fulfilled the inclusion criteria. No significant differences were observed between prehabilitation and control groups for PC, HLS, reoperations, or readmissions. Nonetheless, one study reported more favourable outcomes in PC and reoperations related to previous breast surgery in patients undergoing prehabilitation.

**Conclusions:** Exercise-based prehabilitation may reduce PC and unplanned reoperations associated with prior surgery in patients with BC, although no differences were found for HLS or readmissions compared with usual care. Given the limited number of trials, further research is required to determine the potential benefits of prehabilitation on surgical outcomes in BC.

## Introduction

Breast cancer is the most diagnosed type of cancer worldwide, with an estimation of 2.261.419 diagnoses in 2020 ^1^. According to the World Health Organisation and the American Cancer Society, surgery is the elective and most common treatment for people with breast cancer, followed by radiotherapy, chemotherapy, and hormone therapy, among others ^2,3^. The surgery type varies depending on the disease stage and characteristics ^4^; breast-conserving surgery (i.e., only the tumour and some surrounding tissues are removed) and mastectomy (i.e., the entire breast is removed) being the main surgery types ^3^. Both surgeries may also include additional procedures such as the sentinel lymph node biopsy or axillary lymph node dissection ^3^. Therefore, these are key factors determining surgery’ complexity and, in turn, influencing the patient’s recovery.

Arm swelling and/or numbness, pain, lymphedema, reduced function, or discomfort are some side effects that may affect patients’ recovery and quality of life after breast cancer surgery ^5^. Furthermore, several surgical complications such as bleeding, blood clots, damage to nearby tissues, drug reactions, damage to other organs, pain, or infections may arise during and after surgery ^6,7^. As reported by the GlobalSurg Collaborative et al. ^8^, 5.9% of people with breast cancer undergo major postoperative complications, 36.1% of patients experiencing postoperative complications independently of the patient’ age but influenced by the type of surgery ^9^.

Prehabilitation is a process aimed to improve patients’ physical and psychological health before surgery ^10^. In people with cancer, prehabilitation includes physical activity as an essential component, which may result in improvements in baseline functional levels, reduction of the risk of postoperative complications, enhancement of recovery outcomes and maintenance of health-related quality of life ^10^. Growing evidence supports exercise-based prehabilitation as an indispensable part of the continuum of cancer care ^11^, although there are still some issues that must be clarified to assess the optimal intensity, frequency, and exercise selection ^12–14^ of prehabilitation plans.

Several studies aimed at assessing the effectiveness of exercise interventions as a prehabilitation strategy for people living with cancer have been conducted in recent years. Importantly, clinical trials including patients with different types of cancer showed improvements in surgical prognosis (e.g., shorter length of stay at the hospital and reduced postoperative overall and pulmonary complications), functional capacity (e.g., maximum oxygen consumption or muscular strength), and health-related quality of life ^12–15^. Specifically, exercise-based prehabilitation interventions conducted in people with colorectal cancer have demonstrated to significantly improve functional capacity and reduce complications and length of stay ^16–18^. Similarly, studies including people with gastric cancer have suggested significant enhancements of functional capacity ^19^, and notorious reductions of (i) hospital length of stay ^20^, (ii) postoperative pneumonia ^21^, and (iii) general morbidity ^21^. Moreover, in the case of people with urologic cancer, exercise-based prehabilitation seemed to induce improvements in cardiorespiratory fitness and quality of life ^22^, but the authors reported a lack of sufficient number of studies to draw robust conclusions ^22–25^. Although exercise-based prehabilitation improves upper extremity recovery after surgery ^26^, physical function, psychosocial variables, and quality of life ^27^ in people with breast cancer, whether these interventions may enhance the surgical prognosis remains uncertain.

Therefore, the aim of the present study was to systematically review the existing and available evidence on the effects of exercise-based prehabilitation on postoperative complications, hospital length of stay, reoperations, and hospital readmissions in people with breast cancer undergoing surgery.

## Methods

### Procedures

The present systematic review was conducted following the Preferred Reporting Items for Systematic Reviews and Meta-Analyses (PRISMA) ^28^ and registered in the International Prospective Register for Systematic Reviews (PROSPERO, registration number: <u>CRD42023492902</u>).

### Search strategy and selection of studies

A systematic search was conducted on the electronic databases PubMed and Web of Science from inception to October 2025. The complete search strategy is presented in Appendix A. Studies were selected following the PICOS (population, intervention, comparison, outcomes, and study) strategy. After removing duplicates, eligible articles were screened by title and abstract by two independent reviewers (A.E-S. & P.C.), and any discrepancies were discussed with a third reviewer (A.C-B.). Subsequently, full-text screening was conducted following the same process.

### Eligibility criteria

Eligibility criteria were: (a) randomized and non-randomized controlled trials including people with breast cancer; (b) to develop a prehabilitation intervention including physical activity or exercise training; (c) to assess postoperative complications, length of hospital stay, reoperations, and/or hospital readmissions.

### Data extraction

Data on postoperative complications, length of hospital stay, reoperations, and/or hospital readmissions were obtained. In addition, descriptive characteristics of each study (i.e., authors, year of publication, number of participants, design, intervention type, duration and description, control group intervention, outcomes and main results) were collected. Likewise, characteristics of the participants in each study were compiled (i.e., age, body mass index, and cancer type, and stage). All data were recorded in a previously designed codebook.

### Quality assessment

In order to assess the quality of the included studies, the Quality Assessment Tool for Quantitative Studies ^29^ was used by two independent reviewers, discussing any discrepancy with a third reviewer. In addition, the methodological quality of the systematic review was assessed using the Joanna Briggs Institute Critical Appraisal Tool for Systematic Reviews ^30^ (Appendix B).

## Results

### Studies selection and description

A total of 687 studies were identified in the initial search, of which 112 were removed for duplication. The 575 remaining articles were screened by title and abstract, and 559 were excluded. Of the 16 eligible studies, only two were found to meet the inclusion criteria during the full text review ^31,32^. Figure 1 shows the PRISMA flow diagram of the study selection process.

**Figure 1.**
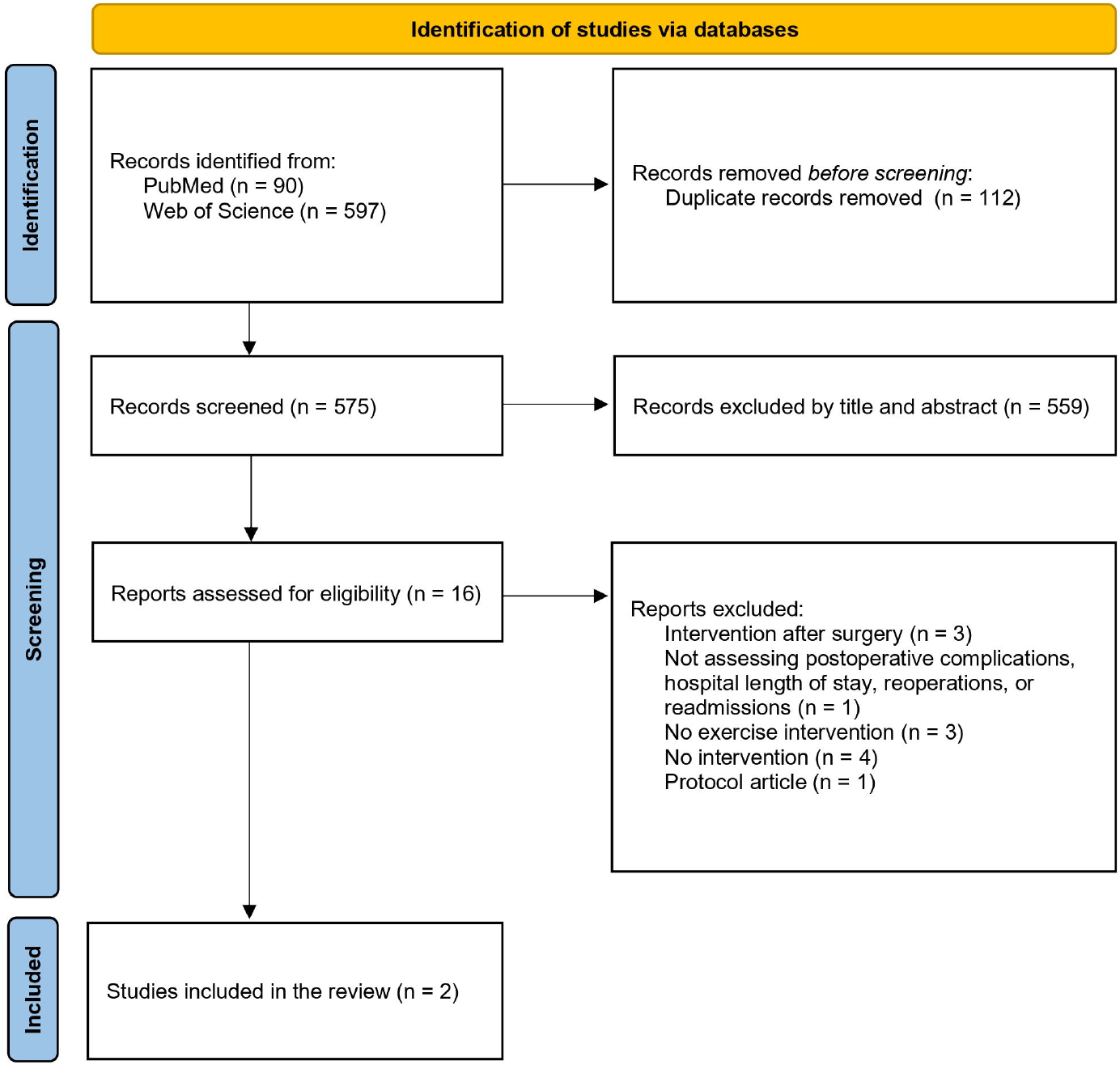
PRISMA flow diagram of the study selection process.

The characteristics of the included studies are presented in Table 1. One study was a randomized controlled trial ^31^ and the other was a non-randomized controlled trial ^32^, including a total of 412 women with breast cancer. Only one study included information about the surgery type and characteristics ^31^. Information about hospital length of stay and complications was reported in both studies 30-days after surgery ^31,32^, while additional information on complications 90-days after surgery was only found in one study ^31^.

**Table 1.**
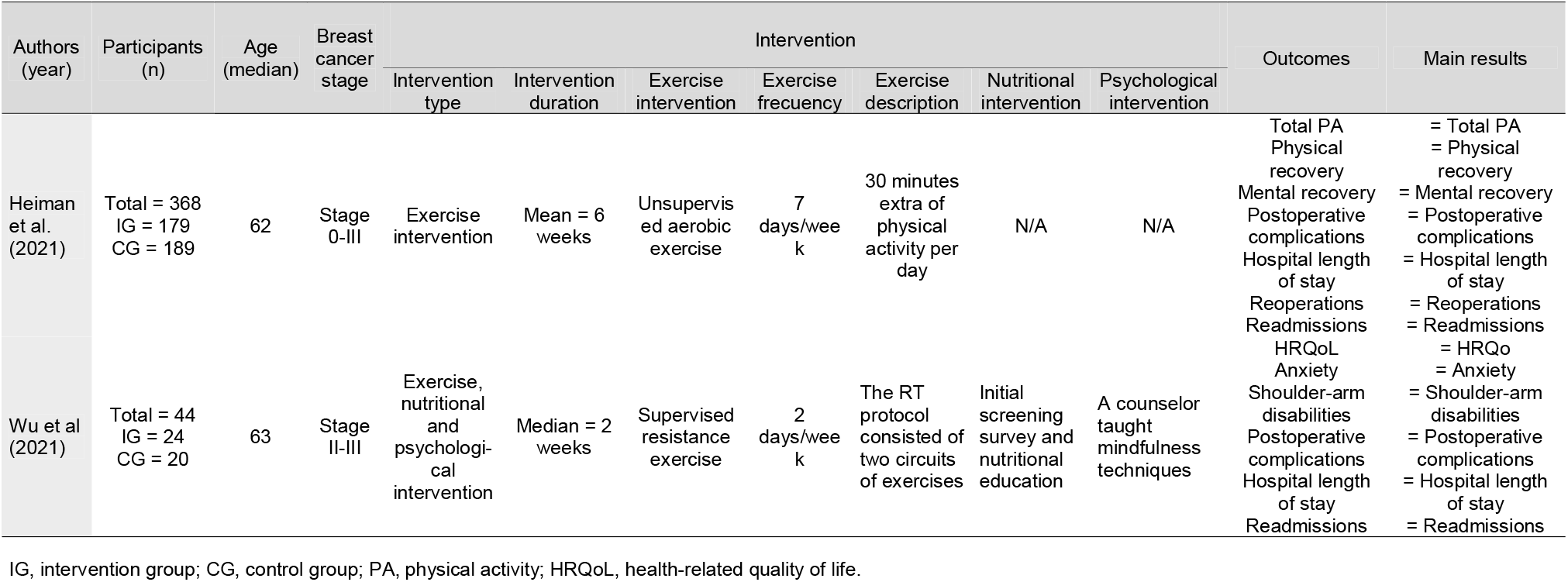
Characteristics of the studies included in the systematic review.

### Postoperative complications

Postoperative complications were reported differently in both studies. Heiman et al. (2021) registered postoperative complications according to the Clavien-Dindo classification and the Comprehensive Complication Index (CCI) at 30 and 90 days after surgery. In contrast, Wu et al. (2020) registered whether any complication that required further hospitalization occurred at 30 days after surgery. Table 2 shows postoperative complications records for both studies. The study by Heiman et al. (2021), reported slightly lower complications rates for all Clavien-Dindo complication grades in the intervention group both at 30 and at 90 days after surgery, although the authors did not report differences between groups. Additionally, lower CCI were observed for the intervention group at each timepoint. No complication was reported by Wu et al. (2020).

**Table 2.** Postoperative complications according to Clavien-Dindo across studies at 30 and 90 days after surgery.

|  | Intervention groups |  | Control groups |  |
| --- | --- | --- | --- | --- |
| Study |  |  |  |  |
| **Heiman et al. (2021)** | **30 days** | **90 days** | **30 days** | **90 days** |
| Grade 0 | 152 (84%) | 137 (77%) | 155 (82%) | 143 (76%) |
| Grade I | 19 (11%) | 23 (13%) | 21 (11%) | 23 (12%) |
| Grade II | 8 (4%) | 17 (10%) | 9 (5%) | 18 (10%) |
| Grade IIIa | 0 (0%) | 0 (0%) | 1 (1%) | 1 (1%) |
| Grade IIIb | 1 (1%) | 2 (1%) | 4 (2%) | 4 (2%) |
| Grade IV | 0 (0%) | 0 (0%) | 0 (0%) | 1 (1%) |
| CCI | 2.3 (0-40.6) | 4.2 (0-57.5) | 3.1 (0-42.7) | 4.7 (0-58.3) |
| **Wu et al. (2021)** | **30 days** | **90 days** | **30 days** | **90 days** |
| Grade 0 | 0 (0%) | N/A | 0 (0%) | N/A |
| Grade I | 0 (0%) | N/A | 0 (0%) | N/A |
| Grade II | 0 (0%) | N/A | 0 (0%) | N/A |
| Grade IIIa | 0 (0%) | N/A | 0 (0%) | N/A |
| Grade IIIb | 0 (0%) | N/A | 0 (0%) | N/A |
| Grade IV | 0 (0%) | N/A | 0 (0%) | N/A |
| CCI | N/A | N/A | N/A | N/A |

CCI, Comprehensive Complication Index.

### Hospital length of stay

Similar values in both intervention and control groups were obtained for the hospital length of stay. Heiman et al. (2021) reported a mean of 1.1 days for participants in the intervention group, while 1.2 days were registered for those in the control group. Similarly, Wu et al. (2020) obtained a median of 2 days for both the intervention and the control groups.

### Reoperations

The number of reoperations was only recorded in the study by Heiman et al. (2021), who reported a total of 16 (eight in each group) unplanned reoperations at 90 days after surgery. Table 3 shows detailed information on reoperations reported by Heiman et al. (2021).

**Table 3.** Reoperations at 90 days after surgery.

|  | Intervention<br>group | Control group |
| --- | --- | --- |
| Heiman et al. (2021) | 90 days | 90 days |
| Due to infection | 1 | 3 |
| Due to bleeding | 2 | 3 |
| Breast reconstruction, revision | 1 | 1 |
| Breast reconstruction, second stage | 0 | 1 |
| Laparoscopic cholecystectomy | 1 | 0 |
| Oncological surgery, head and neck | 1 | 0 |
| Oncological surgery, gynaecology | 1 | 0 |
| Oncological surgery, skin | 0 | 1 |
| Orthopaedic surgery | 1 | 1 |
| Removal of intrauterine contraceptive<br>device | 1 | 0 |
| Central venous catheter | 0 | 1 |

### Readmissions

Unplanned hospital readmissions varied across the studies. Heiman et al. (2021) reported that 8.4% (15) of the participants in the intervention group experienced a hospital readmission within 90 days after surgery, compared with 5.3% (10) in the control group. Wu et al. (2020) reported that no unplanned readmission occurred in the intervention or control groups.

### Quality assessment

Quality assessment revealed moderate ^31^ and weak ^32^ quality of the included studies (Figure 2).

**Figure 2.**
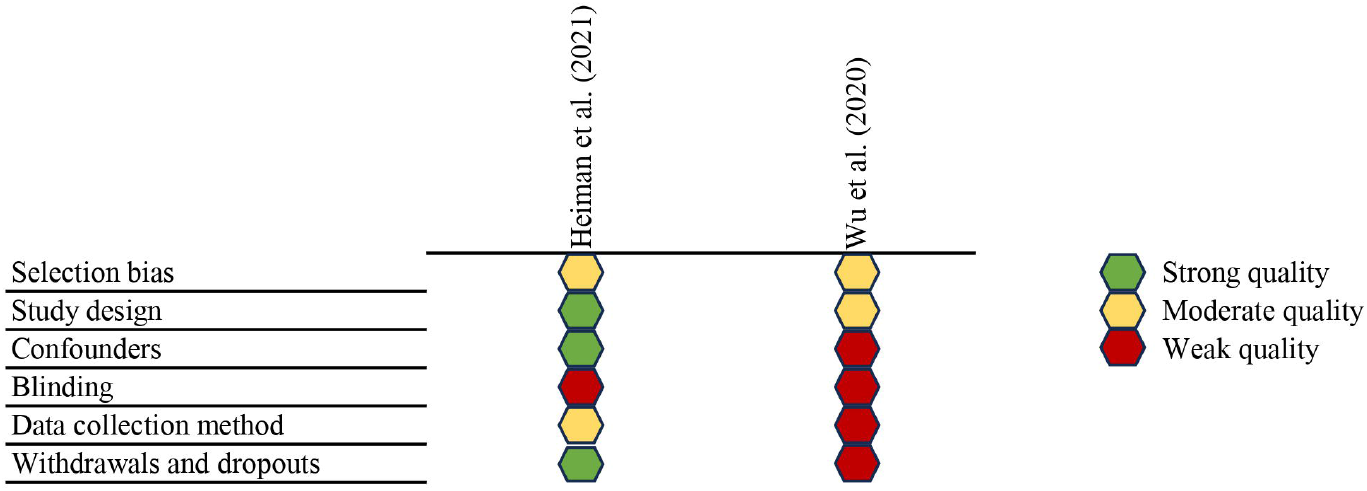
Quality assessment of the included studies.

## Discussion

The present systematic review aimed to revise and update the current knowledge on the effects of exercise-based prehabilitation on postoperative complications, hospital length of stay, reoperations and hospital readmissions in people with breast cancer undergoing surgery. The main findings of this systematic review indicate a significant lack of evidence about the effects of exercise-based prehabilitation on surgical prognosis in people with breast cancer undergoing surgery. The available evidence does not allow to provide robust conclusions, despite a non-significant trend for reducing postoperative complications in one of the only two studies published to date on the topic. The potential clinical relevance of preoperative physical preparation for enhancing surgical prognosis following breast cancer surgery, highlights the urgent need to conduct future rigorous and high-quality randomised trials.

The revised articles hypothesized that engaging in exercise interventions before surgery would reduce postoperative complications, hospital length of stay, reoperations, and readmissions in people with breast cancer, but none of the two studies reported significant differences compared with usual care groups. For instance, the study by Heiman et al. (2021) did not report between-groups differences, but only raw values, suggesting lower postoperative complications for the intervention group, similar values for reoperations in both groups, and lower readmissions for the control group. Nevertheless, the analyses provided do not allow a conclusion on the between-group comparisons. Remarkably, reasons for unplanned reoperations reported by Heiman et al. (2021) were diverse and not all of them were related to previous surgery.

Attending to these reasons, lower reoperations related to previous breast surgery were observed in the intervention group (IG) compared to the control group (CG) (four in IG vs. eight in CG). The study by Wu et al. (2020) reported information on the number of postoperative complications, hospital length of stay and readmissions, although comparisons between groups were not reported. Wu et al. (2020) observed no postoperative complications or readmissions in both the intervention and the control group and similar values for hospital length of stay in the intervention and control groups.

Previous studies have demonstrated that exercise-based prehabilitation programs are effective at improving postoperative complications and hospital length of stay in people with lung, colorectal, gastric, bladder and oesophageal cancer ^12,13,18,21^. Taking this into consideration, the absence of positive effects in people with breast cancer undergoing prehabilitation in the two available studies could be explained by several factors. Firstly, the duration of the interventions □2 weeks before surgery and 4 weeks after surgery in the study by Heiman et al. (2021), and 2 weeks in the study by Wu et al. (2020) □may be insufficient to achieve the potential benefits of exercise-based prehabilitation. Previous literature reporting improvements in surgical prognosis lasted an average of 3-4 weeks ^13,17,18^, although a lack of consensus on the optimal duration is reported by the studies ^12,16,20^. Secondly, the lack of specificity of the exercise-based prehabilitation programs may reduce the potential improvements following prehabilitation interventions. Since breast cancer surgery involves the area comprising the trunk, shoulder and arm, training programs should therefore be focused on strengthening and preparing those musculoskeletal structures for the subsequent surgery ^33^. In the case of the study by Heiman et al. (2021), the intervention consisted of adding 30 minutes of daily physical activity with no further indications, while the exercise intervention by Wu et al. (2020) included 8 resistance exercises involving the full body, with a duration of 30 minutes and a frequency of two sessions per week. Although both programmes may have beneficial effects for people with breast cancer, the specific enhancements for the trunk, shoulder and arm complex may be compromised. Additionally, the studies by Heiman et al. (2021) and Wu et al. (2020) presented a moderate and weak quality, which compromises the validity, replicability, and generalizability of the results.

Remarkably, only the study by Wu et al. (2020), with a sample size of 44, implemented a multicomponent intervention including nutritional and psychological support. The nutritional intervention consisted of advising the participants in terms of consuming adequate amounts of protein and reducing the consumption of processed foods, and the psychological intervention consisted of one session aimed to teach mindfulness to the participants. Multicomponent interventions have demonstrated positive effects on surgical prognosis in people with cancer by reducing postoperative complications and hospital length of stay ^20,21,34^. Therefore, it is noteworthy that only one study implementing prehabilitation in people with breast cancer included nutritional and psychological interventions; this highlights the need of implementing larger studies with higher quality multicomponent interventions in people with breast cancer before surgery ^11^.

Although breast cancer is the most diagnosed cancer type worldwide 1, and leading institutions report that up to 36% of people living with breast cancer undergoing surgery experience some kind of postoperative complications ^8^, there is a lack of well-designed studies assessing the effects of a multicomponent exercise-based prehabilitation on postoperative complications, hospital length of stay, reoperations and hospital readmissions. This systematic review underlines that only two studies with moderate and weak methodological quality implemented a prehabilitation programs in people with breast cancer to improve surgical complications-related outcomes. Given the potential benefits of exercise-based prehabilitation in people with distinct cancer types, assessing the extent to which these interventions may benefit people with breast cancer is of clinical, public health, and economical interest ^35^. Thus, the present study arises the need of implementing exercise-based prehabilitation programmes in people with breast cancer awaiting breast surgery.

This study has several limitations that must be underlined. The limited number of studies included in the review prevents us from drawing robust conclusions. Furthermore, the heterogeneity and the lack of specificity of the training programs limit the comparability of the results from both studies.

## Conclusions

In summary, this systematic review indicates a significant lack of evidence regarding the effects of exercise-based prehabilitation on surgical prognosis in people with breast cancer undergoing surgery. The available evidence does not allow to provide robust conclusions, despite a non-significant trend for reducing postoperative complications in one of the only two studies published to date on the topic. The potential clinical relevance of preoperative physical preparation for enhancing surgical prognosis following breast cancer surgery, highlights the urgent need to conduct future rigorous and high-quality randomised trials.

Breast cancer treatments vary depending on the characteristics of the disease. Therefore, future studies might consider conducting exercise-based prehabilitation interventions including patients with different treatment plans to allow longer prehabilitation interventions. Considering the logistical barriers related to the application of in-person supervised exercise interventions in clinical populations, online interventions may be a feasible alternative for integrating prehabilitation into the breast cancer care continuum.

## Data Availability

All data produced in the present study are available upon reasonable request to the authors

