## Appendices for "Role of exercise prehabilitation on surgical prognosis in people with breast cancer: A Systematic review and call for action"

**Appendix A**

Search terms used in databases.

**MEDLINE (via PubMed)**

(breast[Title/Abstract] OR mammar*[Title/Abstract]) AND (cancer[Title/Abstract] OR oncolog*[Title/Abstract] OR tumor[Title/Abstract])

AND (operat*[Title/Abstract] OR surg*[Title/Abstract] OR resect*[Title/Abstract] OR incision*[Title/Abstract])

AND (prehabilitat*[Title/Abstract] OR pre-habilitat*[Title/Abstract] OR preoperat* [Title/Abstract] OR pre-operat*[Title/Abstract] OR presurg*[Title/Abstract] OR pre-surg*[Title/Abstract] OR before[Title/Abstract])

AND (intervention*[Title/Abstract] OR program*[Title/Abstract] OR trial[Title/Abstract])

AND (exercis*[Title/Abstract] OR resistance[Title/Abstract] OR concurrent[Title/Abstract] OR "physical activit*"[Title/Abstract] OR train*[Title/Abstract] OR lifestyle[Title/Abstract] OR move*[Title/Abstract] OR moving[Title/Abstract] OR sport*[Title/Abstract] OR walk*[Title/Abstract] OR strength*[Title/Abstract] OR endurance[Title/Abstract] OR weightbear*[Title/Abstract] OR weight-bear*[Title/Abstract] OR step*[Title/Abstract] OR ambulat*[Title/Abstract])

AND (CCI[Title/Abstract] OR complicat*[Title/Abstract] OR readmission*[Title/Abstract] OR stay[Title/Abstract] OR hospitalization[Title/Abstract] OR “functional capacity”[Title/Abstract] OR fitness[Title/Abstract] OR physical[Title/Abstract] OR muscular[Title/Abstract] OR “aerobic capacity”[Title/Abstract] OR “maximal oxygen consumption”[Title/Abstract] OR “VO_2_max”[Title/Abstract] OR “VO_2_peak”[Title/Abstract] OR "running speed"[Title/Abstract])

**Web of Science**

TS=(breast OR mammar*) AND TS=(cancer OR oncolog* OR tumor)

AND TS=(operat* OR surg* OR resect* OR incision*)

AND TS=(prehabilitat* OR pre-habilitat* OR preoperat* OR pre-operat* OR presurg* OR pre-surg* OR before)

AND TS=(intervention* OR program* OR trial)

AND TS=(exercis* OR resistance OR concurrent OR "physical activit*" OR train* OR lifestyle OR move* OR moving OR sport* OR walk* OR strength* OR endurance OR weightbear* OR weight-bear* OR step* OR ambulat*)

AND TS= (CCI OR complicat* OR readmission* OR stay OR hospitalization OR “functional capacity” OR fitness OR physical OR muscular OR “aerobic capacity” OR “maximal oxygen consumption” OR “VO_2_max” OR “VO_2_peak” OR "running speed")

**Appendix B**

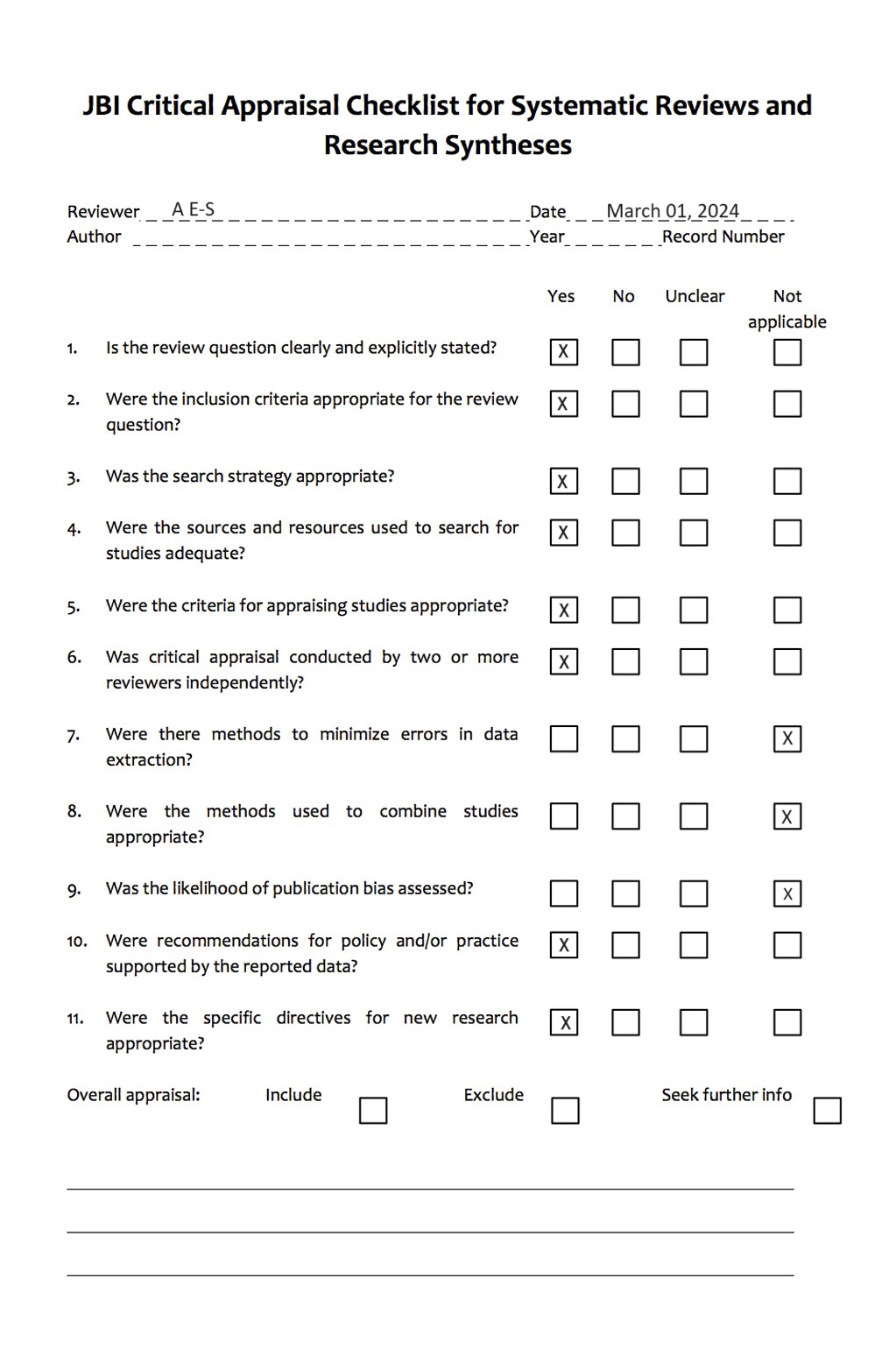
